# Adjunctive Intra-arterial Urokinase after Successful Endovascular Thrombectomy in Patients with Large Vessel Occlusion Stroke (POST-UK): Study protocol of a multicenter, prospective, randomized, open-label, blinded-endpoint trial

**DOI:** 10.1101/2024.08.05.24311528

**Authors:** Chang Liu, Fengli Li, Jiaxing Song, Xu Xu, Jiacheng Huang, Changwei Guo, Weilin Kong, Jie Yang, Xiaolei Shi, Jinfu Ma, Shihai Yang, Zhixi Wang, Shitao Fan, Xiang Liu, Wenzhe Sun, Nizhen Yu, Chengsong Yue, Zhouzhou Peng, Linyu Li, Cheng Huang, Dahong Yang, Duolao Wang, Jeffrey Saver, Thanh N. Nguyen, Raul G. Nogueira, Yangmei Chen, Wenjie Zi

## Abstract

**Background:** Intra-arterial infusion of an adjunctive thrombolytic agent after macrovascular recanalization by endovascular thrombectomy (EVT) was regarded as a promising strategy to promote outcomes of stroke patients. Given the characteristics of urokinase (UK) as an affordable, available, and widely applied medication, especially in eastern countries, this trial aims to assess the safety and efficacy of intra-arterial UK as adjunct to EVT in improving outcomes among patients with anterior large vessel occlusion stroke after excellent to complete reperfusion.

**Methods:** The Adjunctive Intra-arterial Urokinase after Successful Endovascular Thrombectomy in Patients with Large Vessel Occlusion Stroke (POST-UK) trial is a multicenter, prospective, randomized, open-label, blinded-endpoint trial conducted in China. The planned sample size is 498. Those eligible patients with anterior circulation large vessel occlusion stroke and achieving excellent to complete reperfusion by EVT are planned to be consecutively randomized in a 1:1 ratio to the experimental group (a single dose of intra-arterial urokinase) or to standard of care.

**Results:** The primary outcome is a freedom from disability (modified Rankin Scale, mRS, of 0-1) at 90±7 days. The safety outcomes are mortality within 90±7 days and symptomatic intracranial hemorrhage within 48 hours.

**Conclusions:** The POST-UK trial will provide valuable insight of efficacy and safety of intra-arterial UK in patients with large vessel occlusion stroke after achieving excellent to complete reperfusion by EVT.

**Trial registry number:** ChiCTR2200065617 (www.chictr.org.cn).

## Introduction

In recent decades, the annual number of disability and deaths due to ischemic stroke increased substantially worldwide, especially in low-income and lower-middle-income countries^1^. Endovascular thrombectomy (EVT) has achieved level 1A recommendation for select patients in current stroke guidelines.^2^ However, less than 40% of ischemic stroke patients caused by large vessel occlusion (LVO) are disability free at 90 days despite excellent to complete reperfusion (expanded Thrombolysis In Cerebral Infarction [eTICI] scale 2c-3) by EVT.^3^ In addition to baseline infarct volume, the outcomes of LVO patients are determined in part, by the degree of infarct growth after intervention, caused by insufficient reperfusion for microcirculation and microcirculation.^4^ With visible distal emboli, 1-10% of the target artery territory remains hypoperfused among patients with an eTICI 2c technical outcome. In these with eTICI 3 technical outcome, normal cerebral angiogram findings are not necessarily indicative of an effective perfusion of the microvascular bed.^5^ A post-hoc analysis of three trials reported that the prevalence of microcirculatory dysfunction after satisfactory reperfusion of EVT was over 25%.^6^

As visible thrombi in distal arteries and small unobservable thrombi within the microcirculation impaired perfusion status, intra-arterial infusion of an adjunctive thrombolytic agent after EVT was regarded to be a promising strategy to decrease the incidence of disability after macrovascular recanalization by EVT. Recently, the CHOICE trial administered intra-arterial alteplase among patients after successful EVT and demonstrated that this therapeutic strategy could significantly improve the rate of excellent outcome.^5^ However, the CHOICE study was prematurely terminated due to the lack of availability of the placebo, which might have introduced potential bias. Compared with alteplase, urokinase (UK) has received recommendation as an efficient and safe intra-arterial adjunctive medication for EVT in clinical guidelines in east Asia.^7^ Due to the low price and the availability of customized packaging suitable for arterial thrombolysis, UK has gained widespread use, especially in eastern countries. The INFINITY registry reported that administration of intra-arterial UK as adjunct to EVT appeared to achieve better outcomes than intra-arterial alteplase among patients with LVO.^8^ Another multicenter study also showed that adjunctive treatment with intra-arterial UK after EVT was safe and effective in improving angiographic reperfusion among patients with failed or incomplete reperfusion.^9^ However, there is currently no randomized controlled trial evaluating the effect of intra-arterial UK after successful EVT in patients with LVO.

Therefore, given the characteristics of UK as an affordable, available medication especially in eastern countries, we conducted the POST-UK trial, to investigate the efficacy of intra-arterial UK after successful recanalization with respect to functional outcomes as well as its safety including symptomatic intracerebral hemorrhage (SICH) and mortality. We hypothesized that intra-arterial UK after successful EVT would improve the clinical outcomes of patients with LVO as compared with EVT alone.

## Methods

### Design

The POST-UK trial is an investigator-initiated, multicenter, prospective, randomized, clinical trial with open label treatment and blinded endpoint assessment (PROBE). The trial logo is shown in Figure 1. This trial was registered at www.chictr.org.cn (ChiCTR2200065617). The trial was designed in compliance with the Declaration of Helsinki. This study was approved by the Human Research Ethics Committee of The Second Affiliated Hospital of Chongqing Medical University, and all participating centers. This study included 35 stroke centers across China. The key criterion for a qualifying participating center was the performance of at least 60 mechanical thrombectomy procedures with the use of stent-retriever or contact aspiration devices annually, and all participating neuro-interventionalists should have had more than 3 years’ experience and performed at least 50 EVT. The trial flowchart is shown in Figure 2.

**Figure 1.**
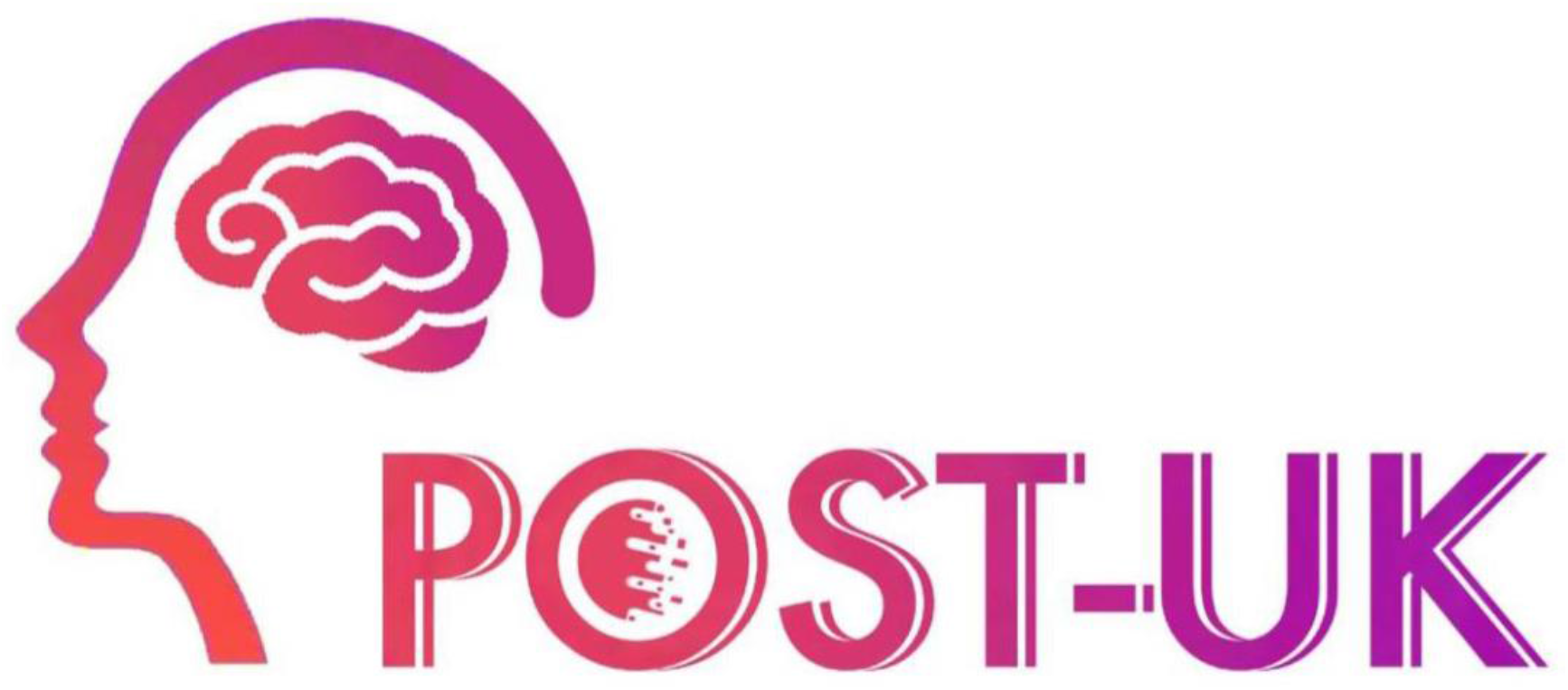
Trial logo

**Figure 2.**
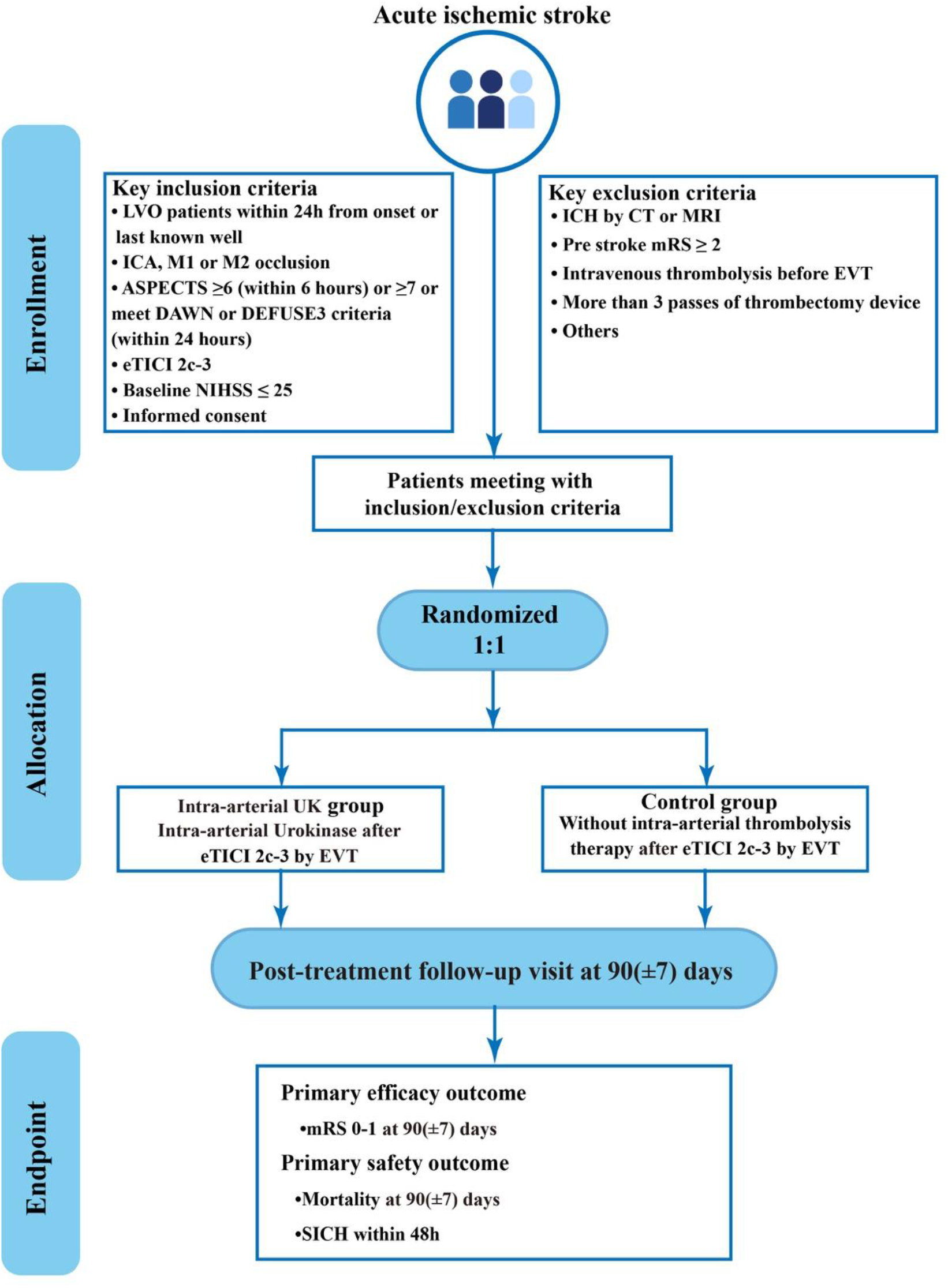
Study flow chart. eTICI, expanded Thrombolysis In Cerebral Infarction; EVT, endovascular thrombectomy; HBC, Heidelberg Bleeding Classification; mRS, modified Rankin Scale; NIHSS, National Institutes of Health Stroke Scale.

### Patient population

The POST-UK trial inclusion criteria are as follows:

1. Age ≥ 18 years;
2. Large vessel occlusion confirmed by CTA/MRA/DSA (intracranial segment of the internal carotid artery (ICA), middle cerebral artery (MCA) M1 or M2 segment);
3. Presenting acute ischemic stroke symptoms within 24 hours of onset (last known well);
4. Baseline National Institutes of Health Stroke Scale (NIHSS) ≤ 25;
5. Baseline Alberta Stroke Program Early CT Score (ASPECTS) ≥ 6 based on non-contrast computed tomography (NCCT) if the time from onset (last known well) was within 6 hours; ASPECTS ≥ 7 or met the Endovascular Therapy Following Imaging Evaluation for Ischemic Stroke (DEFUSE 3) study criteria (infarct volume of less than 70 ml, ratio of volume of 1.8 or more, and an absolute penumbra volume of 15 ml or more) or met the DWI or CTP Assessment with Clinical Mismatch in the Triage of Wake-Up and Late Presenting Strokes Undergoing Neurointervention with Trevo (DAWN) study criteria (I: aged ≥ 80 years, NIHSS ≥ 10, infarct volume < 21 ml; II: aged < 80 years, NIHSS ≥ 10, infarct volume < 31 ml; III: aged < 80 years, NIHSS ≥ 20, infarct volume of 31 to less than 51 ml) within 24 hours;
6. Treated with EVT resulting in an expanded Thrombolysis in Cerebral Infarction (eTICI) score ≥2c
7. Written informed consent (by patients or legally authorized representative).

### Exclusion Criteria

1. Intracranial hemorrhage confirmed by computed tomography (CT) or magnetic resonance imaging (MRI) on admission;
2. Treated with intravenous thrombolysis or contraindication to intravenous thrombolysis (except time to therapy);
3. Prestroke mRS score ≥ 2;
4. Vessel rupture, dissection, or contrast extravasation during the procedure;
5. Procedure time > 90 min;
6. More than 3 passes of thrombectomy device;
7. Females who are pregnant or in lactation;
8. Severe contrast allergy or absolute contraindication to iodinated contrast;
9. Systolic blood pressure > 185 mmHg or diastolic pressure > 110 mmHg, and cannot be controlled by antihypertensive drugs;
10. Known genetic or acquired bleeding diathesis, lack of anticoagulant factors, oral anticoagulants or INR > 1.7;
11. Blood glucose < 2.8 mmol/L (50 mg/dl) or > 22.2 mmol/L (400 mg/dl)
12. Platelets < 90 × 10^9^/L;
13. Bleeding history (such as gastrointestinal and urinary tract bleeding) in the prior one month;
14. Chronic hemodialysis and known severe renal insufficiency (glomerular filtration rate < 30 ml/min or serum creatinine > 220 umol/L [2.5 mg/ dL]);
15. Life expectancy < 6 months;
16. Patient that cannot complete 90-day follow-up;
17. Intracranial aneurysm or arteriovenous malformation;
18. Brain tumors with mass effect on brain imaging;
19. Participating in other clinical trials.

### Randomization

After confirming patient eligibility and obtaining written informed consent, randomization is immediately conducted through a web-based application (**www.jinlingshu.com**). Eligible patients are randomly assigned to the intra-arterial UK group or the control group in a 1:1 ratio using a permuted block randomization method with randomly selected block sizes of 2, 4 or 6. The randomization procedure is web-based and runs on mobile devices or web page platforms.

### Treatment

Both treatment arms received EVT. The modality of EVT including stent retrievers, aspiration, balloon angioplasty, stent deployment, or various combinations of these approaches, are determined as per standard local protocols. After randomization, patients in the UK group receive intra-arterial UK (100,000 IU) through a distal access catheter or microcatheter located proximal to the original occlusion, e.g., ICA, M1, or M2. Patients randomized to the control group will terminate the procedure without additional intra-arterial thrombolysis therapy.

### Efficacy endpoints

The primary endpoint is defined as modified Rankin Scale (mRS) score 0 to 1 at 90±7 days after randomization.

The secondary endpoints are:

1. mRS score 0 to 2 at 90±7 days;
2. level of disability (shift analysis of mRS score) at 90±7 days;
3. the change of the NIHSS score at five to seven days or discharge if earlier from baseline;
4. European Quality Five-Dimension scale score at 90±7 days.

### Safety endpoints

The safety endpoints are the following:

1. mortality at 90±7 days;
2. symptomatic intracranial hemorrhage (SICH) within 48 hours according to the modified Heidelberg Bleeding Classification^10, 11^;
3. any intracranial hemorrhage within 48 hours

### Data and Safety Monitoring Board (DSMB)

An independent DSMB comprising 3 experts (a neurologist, a neuro-interventionist, and a biostatistician independent of the overall study statistician) was established to oversee the overall conduct of this trial. Members of the DSMB will not participate in the trial or be affiliated with the study sponsors. The DSMB will meet annually and monitor trial progress. The experts in DSMB will review the incidence of symptomatic intracranial hemorrhage, and serious adverse events to ensure the safety of enrolled patients.

### Sample size estimate

Based on preceding RESCUE BT trial, we assumed that the proportion of excellent functional outcome (mRS 0-1) was 32.8% in the control group^12^. With a 18.6% absolute difference of the mRS 0-1 in the CHOICE trial, we conservatively estimated a 13% difference between the control group and the treatment group, which indicated the proportion of mRS 0-1 in the treatment group would be 45.8%^12, 13^. To demonstrate a 13% absolute difference with a type-1 error alpha of 0.05 (two-tailed) with a power of 83%, a sample size of 472 patients would be needed (236 patients per group). To take into account a 5% attrition rate, a total of 498 patients was required (249 per group). This sample size calculation was performed on PASS (NCSS, LLC, Kaysville, USA) version 15.0.

### Statistical analysis

The primary endpoint will be analyzed using Generalized Linear Model (GLM), which compares the proportion of patients with an excellent outcome (mRS 0–1) at 90 days between the two treatment groups and generates the estimate of risk ratio and its 95% confidence interval (CI) as the measurement of treatment effect. The estimate will be adjusted for age, baseline NIHSS, baseline ASPECTS, occlusion sites, and time from onset to randomization. The secondary outcomes and safety outcomes will be analyzed using GLMs or assumption-free methods as applicable^14^, with the same adjusting method for the covariates listed above used for the primary outcome analysis. Missing values for baseline characteristics and outcomes will be reported. Adjusted and unadjusted estimates of treatment effects with corresponding 95% CIs will be reported. All analyses will use a 5% two-sided level of significance. The primary analyses will be based on the intention-to-treat principle. The per-protocol analyses will also be performed as supplemental analysis. Statistical analyses will be performed using SAS 9.4 and R 4.3.0. The trial results will be reported following the CONSORT guidelines for reporting randomized trials. All analyses will be detailed in the statistical analysis plan which will be finalized before the unblinding of the study data.

## Discussion

With the increase of stroke-related disability and death worldwide, especially in lower-income and lower-middle-income countries, exploring cost-effective strategies to improve outcomes of LVO is needed.^15^ Hence, the multicenter, prospective, randomized clinical trial of POST-UK aims to investigate whether the a low-dose widely available clinical thrombolysis agent, UK, confers benefit as adjunctive therapy to EVT among patients with LVO who achieved excellent to complete reperfusion by EVT.

Approved by the National Medical Products Administration in China, UK has been recommended in the latest Chinese stroke guidelines for intravenous thrombolysis use in acute ischemic stroke patients.^16, 17^ At less than one-tenth the cost of alteplase, UK has gained widespread use, especially in the lower-income and lower-middle-income countries.^18^ As only a small dose of thrombolytic agent is required for intra-arterial thrombolysis, the low dose packaging of UK facilitates its application in EVT.^19^ Preclinical evidence also showed that UK upregulates blood brain barrier tight junctions and has a favorable effect on matrix metalloproteinases, potentially lowering the risk of bleeding.^20^ Compared with intravenous alteplase, a meta-analysis reported that intravenous UK was as safe and effective in the treatment of acute ischemic stroke.^18^ For intra-arterial thrombolysis, a study comprising 311 patients showed a numerically lower rate of sICH and numerically higher rate of favorable outcomes after intra-arterial UK than alteplase after EVT.^8^ However, up to now, there has been no clinical trial dedicated to the use of intra-arterial UK in the management of patients after successful reperfusion with LVO.

The application of 100,000 IU of UK in our trial was based on a review of prior literature. Pilot studies reported that 100,000 IU of UK could improve the prognosis among patients with LVO, and it was also effective in preventing new blood clots^21-23^. Moreover, the MELT Japan trial showed that intra-arterial dose ranges of UK between 120,000 IU to 600,000 IU for fibrinolysis of middle cerebral artery occlusion was safe. We selected a lower dose of intra-arterial UK because in contrast to MELT, patients in POST-UK already had successful recanalization.^24^ As the mean dose for intra-arterial alteplase in the CHOICE trial was 16.65 mg, our intra-arterial UK dose of 100,000 IU was proportional to the alteplase dose.^13, 24^

To minimize the risk of intracranial hemorrhage, we included patients after imaging screening and we excluded patients who had received IVT.^25, 26^ We excluded patients with more than 3 passes of a thrombectomy device as this can be associated with higher risk of endothelial injury with greater instrumentation of the vessel.^27^ Based on the subgroup analysis from the CHOICE trial, which did not show an improvement of outcomes among patients with eTICI 2b with intra-arterial thrombolysis, only patients with eTICI of 2c to 3 were enrolled in the current trial due to their lower frequency of sICH compared with patients with eTICI 2b.^13^

We acknowledge limitations in the current trial. The treatment assignment is open-label, which may lead to potential assessment bias in the study outcomes. To mitigate this limitation, independent and blinded adjudication of the primary outcome assessment at 90 days was utilized. Independent core lab adjudication of imaging results with readers blinded to treatment assignment will also be conducted. The sample size of our trial was based on the results of a phase IIb randomized trial of modest sample size, with a risk difference of 18.6%.^13^ The present trial was designed with the risk difference of 13% in the primary outcome. Though our treatment effect calculation may have been overestimated, the sample size of this study is by far the largest of similar research to date. ^28^

## Conclusion

The POST-UK trial will provide pivotal data to assess the efficacy and safety of intra-arterial UK as adjunct to excellent to complete reperfusion by EVT in patients with anterior circulation LVO.

## Data Availability

The data that support the findings of this study are available on request from the corresponding author, upon reasonable request.

